# Prevalence and determinants of undernutrition among adults living with HIV in Ethiopia: A systematic review and meta-analysis

**DOI:** 10.1101/2025.08.01.25332678

**Authors:** Amare Admasu, Takele Tadesse, Amene Abebe, Eskinder Wolka

**Affiliations:** Wolaita Sodo University College of Health Science and Medicine, Wolaita, Addis Ababa, Ethiopia

**Keywords:** Undernutrition, food insecurity, opportunistic infections, People Living with HIV

## Abstract

**Background:** Undernutrition among HIV-positive adults in Ethiopia highlights the cycle of cause-and-effect relationships between undernutrition and HIV infection.

**Aim:** The study aimed to assess the prevalence of undernutrition and its determinants among HIV-positive adults in Ethiopia.

**Methods:** Intensive searches were carried out utilizing PubMed, EMBASE (Elsevier), Cochrane, and and other electronic databases such as Science Direct, African Journal Online (AJOL), Google Scholar, and gray literature.

**Data synthesis/findings:** A total sample size of 5,648 and a total number of undernourished individuals of 1,474 from 11 articles met the inclusion criteria. The study found that the pooled prevalence of 26.70% (95% CI: 21.31%, 32.10%) of HIV-positive patients in Ethiopia are undernourished. Factors determining undernutrition include having opportunistic infections (OR: 3.496, 95% CI 1.776-5.217), being at an advanced WHO clinical stage II or above (OR: 2.916, 95% CI 1.088-4.744), having a cluster differentiation (CD4) count below 200 cells (OR: 3.099, 95% CI 1.418-4.779), and food insecurity (OR: 3.352, 95% CI 1.418-5.287).

**Conclusion:** The systematic review and meta-analysis found that Ethiopia’s HIV-positive population faces high undernutrition rates in comparison to studies done in sub-Saharan Africa. Opportunistic infections, advanced WHO clinical disease stage, CD4 counts below 200 cells/mm³, and food insecurity were identified as statistically significant factors determining the high prevalence of undernutrition. This suggests a cyclic link between undernutrition and health outcomes. The meta-analysis identifies factors influencing undernutrition in Ethiopia’s HIV-positive population, but more research is needed to determine the efficacy of interventions and address root causes.

## 1. Introduction

### Background

Undernutrition is a condition in which an individual’s body function is impaired and they are unable to maintain normal body performance (<u>1</u>). It is a combination of acute malnutrition (bilateral pitting edema and/or wasting) and micronutrient deficiencies (<u>2</u>, <u>3</u>). Undernutrition and the Human Immunodeficiency Virus(HIV) infection are related and increase one another’s effects in a vicious cycle (<u>4</u>). Undernutrition (underweight) reflects an individual with a BMI of less than 18.5kg/m^2^ (<u>5</u>). The severity of undernutrition is further classified as severe (BMI <16 kg/m^2^), moderate (BMI 16–16.99 kg/m^2^), or mild (BMI 17–18.48 kg/m^2^) (<u>6</u>).

In 2022, 39 million people worldwide were infected with HIV (<u>7</u>). Undernutrition is both a cause and a consequence of HIV/AIDS (<u>8</u>). In 2018, Africa had 256.1 million cases of undernutrition, making it the world’s third most common after Asia (<u>9</u>). In the same period, approximately 70% of the 37.9 million people living with HIV (PLHIV) worldwide resided in Sub-Saharan Africa (SSA) (<u>10</u>).

According to several regional studies, the SSA has the highest rate of undernutrition among PLHIV globally. Studies conducted in various sections of the SSA revealed 43.3% in Nigeria (<u>11</u>), 19.4% in Tanzania (<u>12</u>), 19.2% in Senegal (<u>13</u>), 13.8% in Ghana (<u>14</u>), 10.28% in Uganda (<u>15</u>), and 10% in Zimbabwe (<u>16</u>). Ethiopia is one of the SSA countries most affected by the HIV/malnutrition epidemic. Undernutrition was pervasive across the country, with 63.5% in Bahir Dar(<u>17</u>), 31.2% in Debre Markos (<u>18</u>), 34% and 30% in the Oromia region (<u>19</u>, <u>20</u>), 60% in the Benishangul Gumuz Regional State (<u>21</u>), 31.2% and 25.5% in South and Central Ethiopia (<u>22</u>, <u>23</u>).

To the level of our knowledge, this systematic review and meta-analysis is the first review to clearly report the pooled odds ratio of the determinant factors of undernutrition at the national level. We found only one systematic review done in the country, which reported the pooled prevalence of undernutrition but not the pooled odds ratio of determinant factors by Alebel, A., et al. (<u>24</u>). Therefore, the systematic review and meta-analysis aim to comprehensively assess the pooled prevalence and determinants of undernutrition among adults living with HIV (PLHIV) in Ethiopia. Understanding these determinants is essential for designing effective nutrition and health programs. By identifying the key factors contributing to undernutrition, stakeholders can tailor interventions to meet the specific needs of this population. Furthermore, such insights can inform the development of long-term strategies that enhance nutritional outcomes and overall well-being for PLHIV.

## 2. Methods

### 2.1. Reporting and protocol registration

The methodology and reporting for this review were guided by the Preferred Reporting Items for Systematic Reviews and Meta-Analyses (PRISMA; <u>S1 Table 1</u>) (<u>25</u>). The review protocol was prospectively registered in the international prospective register of systematic reviews, PROSPERO, with ID: CRD42023403023. This review protocol is available at https://www.crd.york.ac.uk/PROSPERO/view/CRD42023403023 ID=CRD42023403023. This evaluation did not require ethics board approval because it focused on summarizing publicly available study findings. Furthermore, data from previously published studies with informed consent received from the primary investigators were downloaded and examined. However, the approaches were sequential, beginning with a literature search and ending with a synthesis of the results.

### 2.2. Search strategies

We looked for published evidence and a wide range of literature, including peer-reviewed articles and grey literature. Intensive searches were carried out utilizing PubMed, EMBASE (Elsevier), Cochrane, and other electronic databases such as Science Direct, African Journal Online (AJOL), Google Scholar, and gray literature. In the PubMed advanced search, we used MeSH terms such as "undernutrition," "malnutrition," "AIDS," "HIV," "adults," "adolescents," "young adults," "acute malnutrition," "recovery," "nutritional deficiency," "nutritional status," "underweight," "stunting," "wasting," and "Ethiopia," as well as Boolean operators like "OR," "NOT," or "AND." The PubMed online database’s search method focuses on detecting undernutrition and related determinants among adults living with HIV through 2020-2025.

#### Research questions

- What is the prevalence of undernutrition among adults living with HIV in Ethiopia?
- What are the major determinants of undernutrition among adults living with HIV in Ethiopia?

### 2.3. Eligibility criteria

#### Inclusion criteria

The studies were included in the review if they fulfill the following criteria:

**A. Study design**: observational studies (including case-control, cohort and cross-sectional studies).
**B. Participants:** adult people living with HIV (PLHIV) (age 15 and above)
**C. Outcome measurments**: prevalence of undernutrition and its determinant factors
**D.** The primary goal of this research is to determine the pooled prevalence of undernutrition among persons with HIV in Ethiopia. It also identified risk factors for undernutrition. Any measure of undernutrition among HIV-infected people was included. Articles published within the last five years, from 2020 to 2025.
**E. Comparator:** well-nourished adults living with HIV
**F. Growth reference:** WHO growth standards. The rationale for using WHO growth standards is to provide a globally consistent and reliable reference point for assessing nutritional status, allowing for accurate detection of malnutrition across diverse populations.
**G.** All articles published only in the English language and gray literature were also included.

#### Exclusion criteria

Studies were excluded if they included:

A. Studies on micronutrient deficiency.
B. Smaller studies increase the risk of bias; studies with a sample size of less than 100 were excluded. Smaller studies have lower statistical power, which can lead to potentially unreliable results and wider confidence intervals, making it difficult to accurately estimate the true effect size and increasing the risk of bias (<u>26</u>).
C. Has a poor quality score (<6/10). This to ensure the overall reliability and validity of the findings by only including studies with a sufficiently rigorous methodology, thereby reducing the risk of bias introduced by including low-quality studies, which could skew the results and lead to incorrect conclusions (<u>27</u>).
D. Reviews, experimental studies, letters, commentaries and editorials.
E. The research does not have the full text in the web address.
F. Unpublished studies
G. Dupilication studies were also checked and removed before the analysis process started.

### 2.3. Screening

Three researchers (A.A, AA, and EW) individually selected studies that matched the inclusion criteria. The titles and abstracts were examined for inclusion, and duplicate citations were removed. After reviewing titles and abstracts against the inclusion criteria outlined above, the researchers collected the complete texts of potentially eligible papers. To ensure that qualifying research was included, the researchers ran the entire text through a systematic and pre-tested approach. In the event of conflicts, they were resolved through consensus among the researchers or by consulting a third author (TT). The researchers used EndNote software (version X7) to import papers from electronic databases and manage citations.

### 2.4. Data extraction

Three reviewers (A.A, AA, and EW) worked individually to gather data from the included studies. A predefined data extraction format on a Microsoft Excel spreadsheet was used to collect information on the name of the author or authors, the year of publication, the study design, the place of the study, the region of the study, the sample size, the age range of study participants, the sex of study participants, the prevalence of undernutrition, and pertinent risk factors or determinant factors of undernutrition (<u>S2A Table 2</u>).

#### Outcome measurement

Anthropometric indicators, namely stunting, underweight, wasting, or thinness, were used based on WHO growth standards (<u>28</u>). Undernutrition (underweight) refers to an individual with a BMI of less than 18.5 kg/m^2^. The severity of undernutrition is further classified as severe (BMI 16 kg/m^2^), moderate (BMI 16–16.99 kg/m^2^), or mild (BMI 17–18.48 kg/m^2^) (<u>6</u>).

### 2.5. Assessment of methodological quality and risk of bias

The reviewers (A.A, TT, AA, and EW) assessed the quality of studies by adopting the Newcastle-Ottawa Quality Assessment Scale (NOS) (<u>27</u>). The first section focused on the methodological quality of each original study, such as objectives, sample size, and sampling technique. Studies with >6 scores were included in the review and meta-analysis of prevalence. All authors independently assessed the quality of the mutual consensus. Any disagreements were resolved by discussion with an independent reviewer. Similarly, in the case of determinant factors, each determinant or factor associated with the outcome variable was critically assessed. Similar cut-off points were employed for all prevalence studies of undernutrition (<u>S2B Table 2</u>).

### 2.6. Data extraction

Using a data extraction template created on Microsoft Excel Spreadsheet 2010, three reviewers (A.A, AA, and EW) independently extracted data from the included studies. The primary author’s name, study region, study region, study period, sample size, estimates of undernutrition prevalence, response rate, and factors were all recorded.

### 2.7. Data analysis and assessment of publication bias

Three reviewers (A.A, AA, and EW) evaluated the quality of the publications included in the review using the Joanna Briggs Institute (JBI) Critical Appraisal Checklist for Studies Reporting Prevalence Data (<u>29</u>). Based on nine criteria, the checklist evaluates the methodological quality of prevalence studies. Yes, no, uncertain, or not relevant were possible options.

### 2.8. Data processing and analysis

The data for this review was collected on a standard Microsoft Excel form and analyzed using STATA version 14.0 software. We used to combine the results of relevant studies. The pooled prevalence of undernutrition and the pooled odds ratio of associated factors were calculated using a random-effects model with a 95% confidence interval. To assess study heterogeneity, Cochran’s Q chi-squared test and Higgins and Thompson’s I² statistic were used. The cut-off I^2^ values of 0%, 25%, 50%, and 75% were used to represent no, low, moderate, and high levels of heterogeneity (<u>30</u>). Publication bias was evaluated by visually inspecting a funnel plot and using the Egger test, with significance determined at a p-value below 0.05. Subgroup analyses were performed according to geography (Amhara, Southern Nations, Nationalities, and Peoples’ Region (SNNPR), Oromia, Addis Abeba, Tigray, and Harari region), study setting (community or institution-based), author and publication year, sample size, cases, and gender. The robustness of the combined data was assessed using a sensitivity analysis, in which each study was eliminated individually to determine its influence.

### 2.9. Heteroginity across the studies

Heterogeneity among reported proportions was assessed by computing p-values of Cochrane Q-statistics and the I² test (<u>31</u>). In this study, substantial variation in results was found among the included studies (I^2^ = 95.7%, p < 0.001). As a result, a random-effects meta-analysis model was used to estimate the DerSimonian and Laird’s pooled effects.

## 3. Results

### 3.1. Study selection

The process of searching and retrieving articles is illustrated in Figure 1. The following databases were utilized for the article search: PubMed with 5,817 articles, ScienceDirect with 1,721, EMBASE (Elsevier) with 10, African Journals Online (AJOL) with 3,550, Cochrane with 27, and Google Scholar with 5,930. A total of 17,055 articles were discovered. After manually filtering for duplicates using EndNote software, 986 articles were deleted. We then screened the remaining 16,069 articles by title and removed 16,005 that were irrelevant. Out of the 64 papers evaluated for full text and abstract, 12 research were removed because they were conducted outside of Ethiopia. Furthermore, 19 papers were deemed unrelated to the study, four studies were removed because they featured non-HIV-positive populations, and one study focused entirely on elderly individuals. This left us with 28 studies, 17 of which were removed due to characteristics such as research population, setting, result, design, and duplication. In the end, 11 papers were included in the final analysis, offering a thorough examination of the research field (<u>S1 Figure 1</u>).

**Figure 1:**
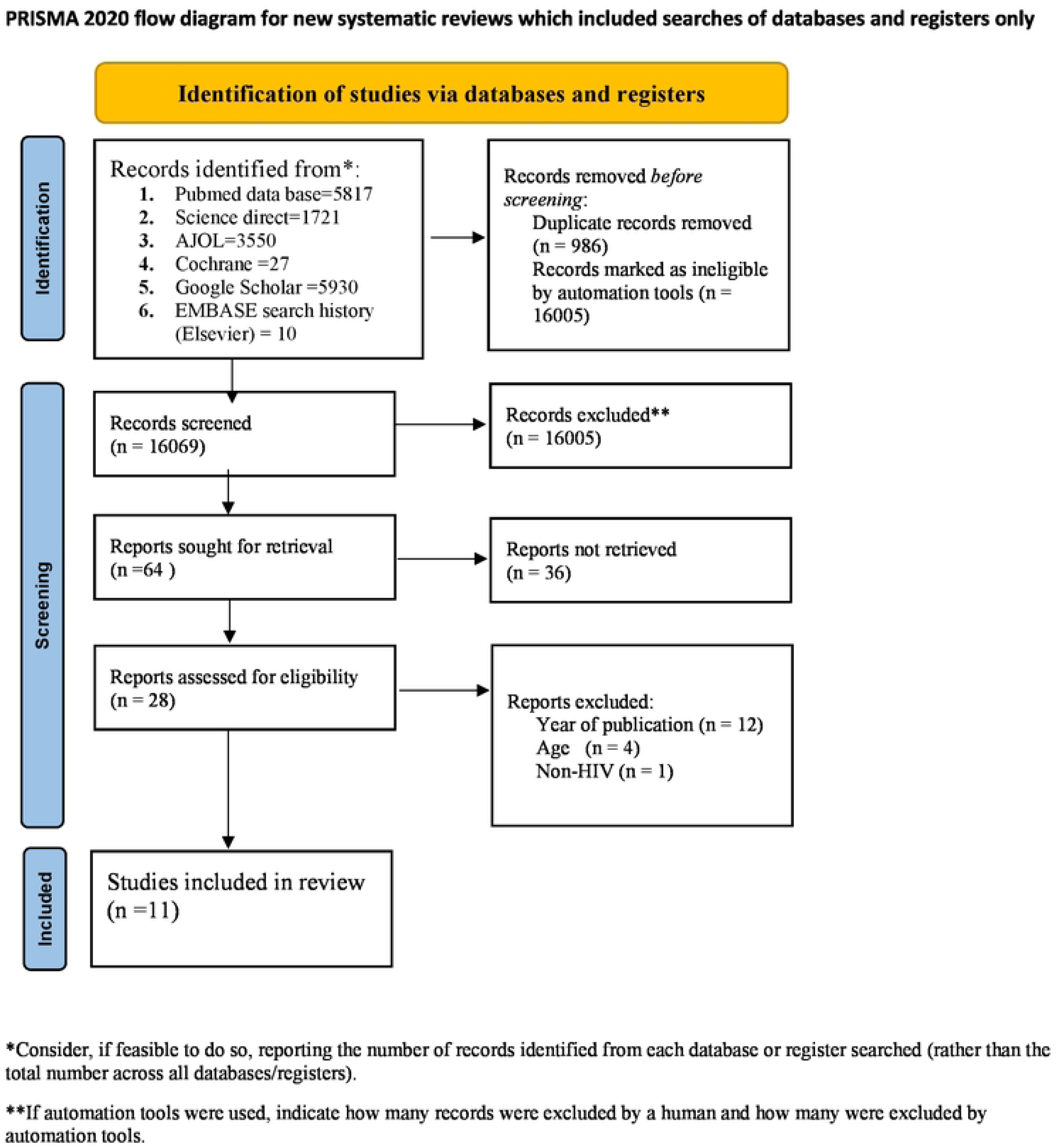
Flow diagram of Systematic review and meta-analysis chart showing the sequence of study selection on undernutrition among adult PLWHIV in Ethiopia, 2025. Source: Page MJ, et al. BMJ 2021;372:n71. doi: 10.1136/bmj.n71. This work is licensed under CC BY 4.0. To view a copy of this license, visit https://creativecommons.org/licenses/by/4.0/

### 3.2. Characteristics of included studies

The main characteristics of all included studies presented in data abstraction summary sheet <u>S2A</u> and quality assessment result <u>S2B</u> Tables. Overall, we got 11 studies that fulfilled all eligibility criteria and remained for systematic review and meta-analysis (<u>19</u>, <u>22</u>, <u>32</u>–<u>40</u>) (<u>S1 Figure 1</u>). These studies were cross-sectional and took place in seven regions of Ethiopia from 2020 to 2025. A total sample size of 5,648 and a total number of undernourished individuals of 1,474. Of the total participants, 3,151 (55.8%) were female, and their counterparts were male. Regional distribution of the included studies: four studies from each of the Oromia regions(<u>19</u>, <u>32</u>, <u>35</u>, <u>40</u>), three from the SNNPR (<u>22</u>, <u>37</u>, <u>39</u>), two from Addis Ababa (<u>34</u>, <u>38</u>), one from Harari (<u>36</u>), and Tigray regions (<u>33</u>) ( <u>S3A Table 3</u>).

### 3.3. Pooled prevalence of undernutrition among PLHIV in Ethiopia

The prevalence of undernutrition among PLHIV was ranging 42.9% in Tigray region (<u>33</u>) to 16% in Bench Maji Zone South Ethiopia region (<u>37</u>). In accordance with the random effect model analysis, the pooled prevalence of undernutrition among PLHIV in Ethiopia was 26.702 %, with a 95% CI (21.306-32.098) with the hetrogenity index (I^2^) of 95.7% (P-value <0.001), confirming significant variability among the studies (<u>S2</u> Figure 2).

**Figure 2:**
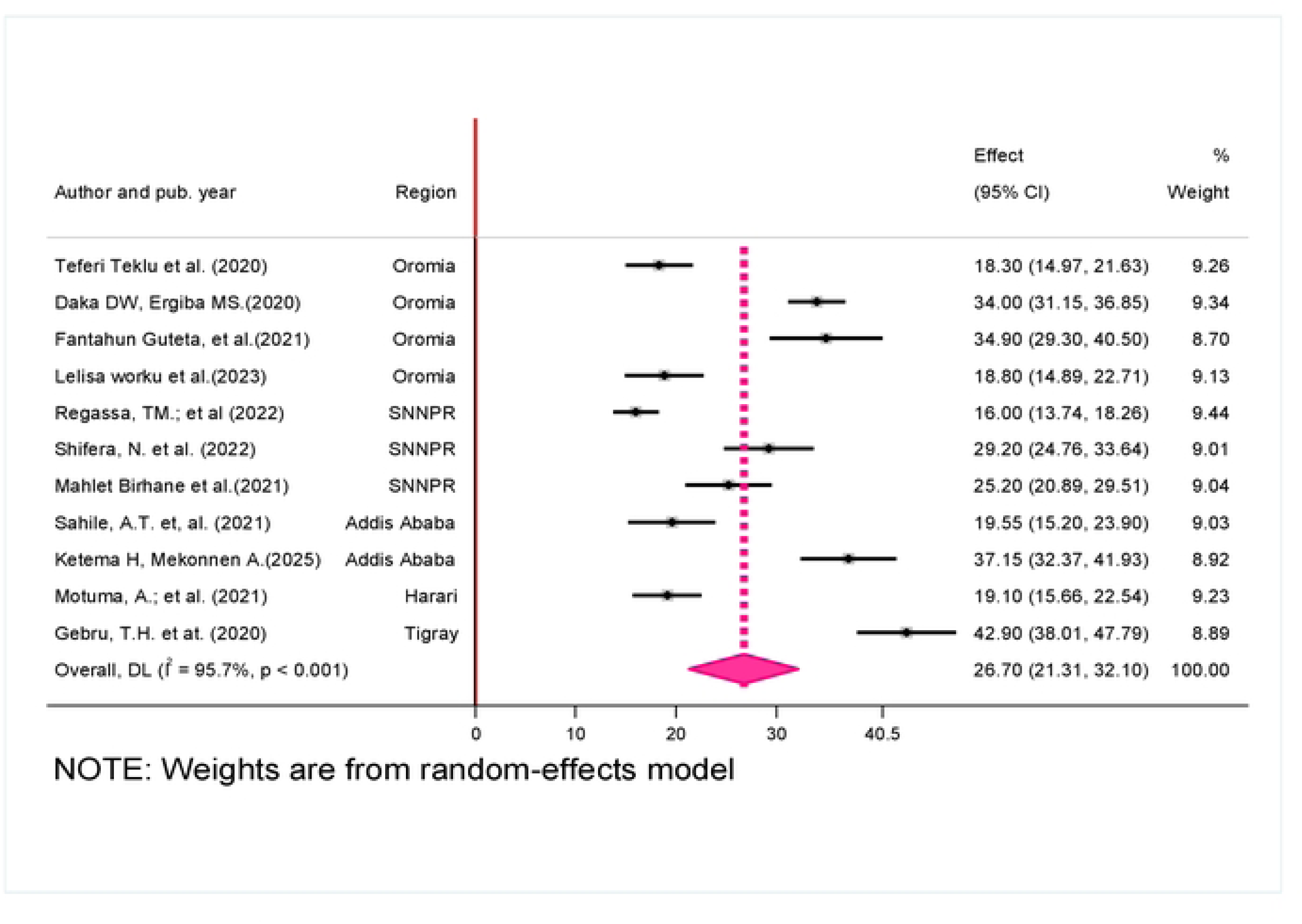
The forest plot presenting pooled prevalence of undernutrition using random effects, effect size,95¾ CI, and weights of each study among PLHIV in Ethiopia, 2025.

### 3.4. Subgroup analysis

The S4A table provides the findings of the subgroup analysis on the pooled prevalence of undernutrition, examining variations among subgroups. The Tigry region had the highest undernutrition prevalence 42.9 (95% CI 38.013-47.787), while the Harari region had the lowest prevalence 19.100 (95% CI 15.661-22.539) (<u>S3A Table 3</u>). There were 7 studies with a sample size of 500 or less, and the highest prevalence of undernutrition was 29.583 (95% CI 22.796-36.370). In terms of data collecting strategies, studies that used interviewer-administered questionnaires had the highest prevalence 37.150 (95% CI 32.373-41.927), compared to those that utilized dichotomous approaches 24.685 (95% CI 19.386-29.984). The difference was statistically significant (p = 0.002) ( <u>S3</u> Figure 3).

**Figure 3:**
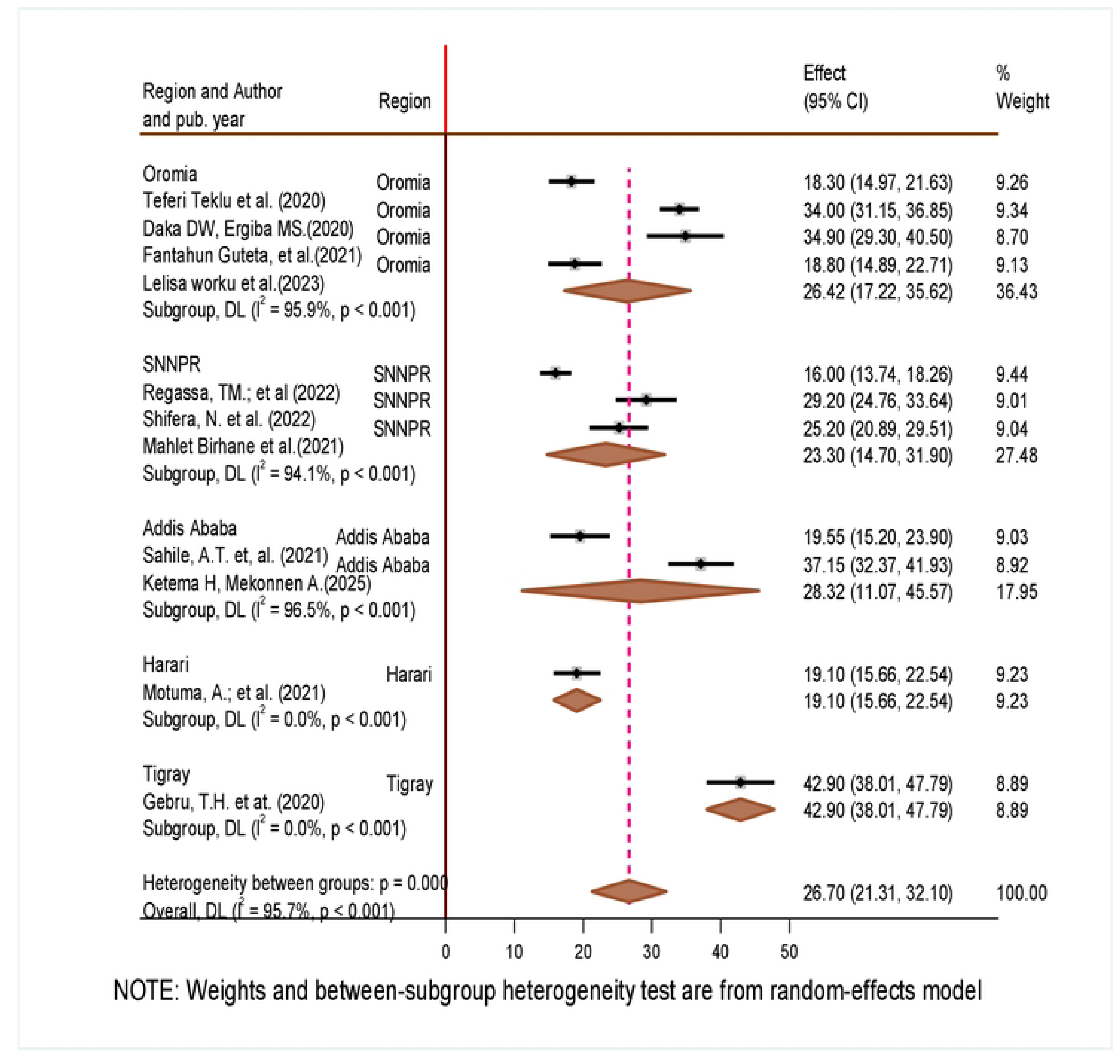
Sub-group analysis of studies from regions included in meta-analysis on the prevalence of undernutrition among PLHIV in Ethiopia, 2025.

### 3.5. Publication bias

The funnel plot and the Eggers test were employed to determine the presence of publication bias in the meta-analysis. The funnel plot of the papers used in the meta-analysis reveals the prevalence of undernutrition among PLHIV in Ethiopia. The Egger test (p = 0.062) offered additional evidence for the absence of publication bias, demonstrating a lack of publication bias in the analysis (<u>S4</u> Figure 4).

**Figure 4:**
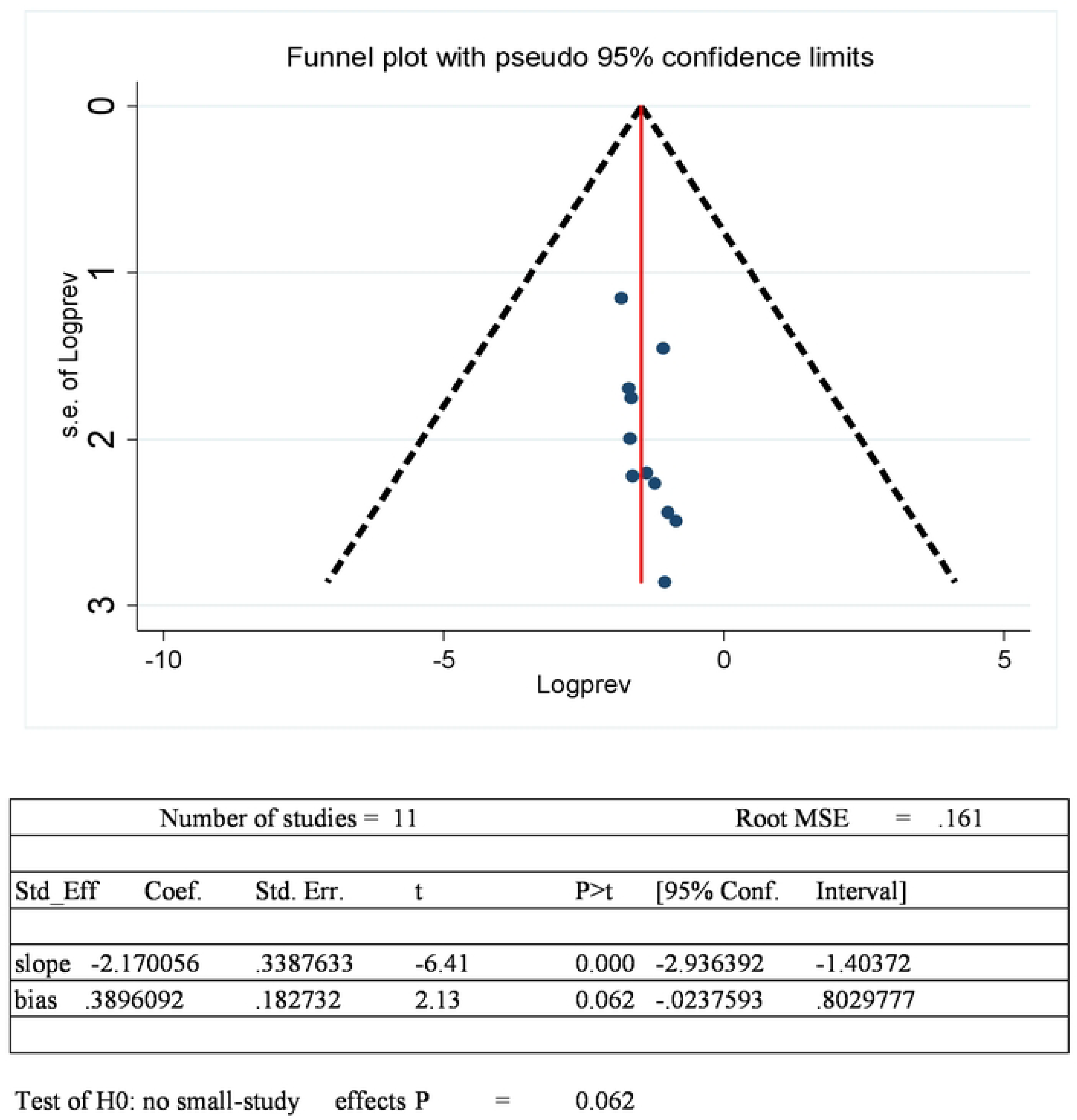
Funnel plot and egger test for pooled prevalence of undernutrition among PLHIV in Ethiopia.

### 3.6. Sensitivity analysysis

To evaluate the diversity in undernutrition findings among PLHIV, the study performed a leave-one-out sensitivity analysis. Excluding each study from the analysis had no significant effect on the average estimated prevalence of undernutrition, which remained within the 95% confidence interval of the total average. As a result, the presence or absence of a single study had no effect on the average prevalence of undernutrition among people living with HIV. Furthermore, the sensitivity findings show that when each study was excluded, the average prevalence of undernutrition varied between 26.70% (95% CI: 7.66, 7.66, 68.51) (<u>S4 Table 4</u>).

### 3.7. Meta-regression

A meta-regression analysis with a random-effects model was used to investigate potential sources of heterogeneity among the included studies. Regions, years of publication, sample size, quality scores, and data collection tools were all considered study-level variables. The meta-regression analysis found no significant relationships between study characteristics (such as regions, year of publication, sample size, quality scores, and data collection tools) and observed results (all statistical values were greater than 0.05). As a result, the differences reported in the studies were not explained by the parameters examined in the analysis. The lack of significant moderators could be attributed to the meta-regression’s small sample size, which weakens statistical power. Other unmeasured variables, such as socioeconomic status throughout the study regions, may contribute to the observed heterogeneity (<u>S5 Table 5</u>).

### 3.8. Determinant factors of undernutrition among adults (PLHIV)

Some of the factors associated with undernutrition in PLHIV were quantitatively pooled in this systematic review and meta-analysis, whereas others were not due to inconsistencies in the definition of independent and dependent variables. As a result, this meta-analysis covers determinants that were repeatedly reported in many original researches. Five of eleven studies found that opportunistic infections (OIs) were a significant predictor of undernutrition in PLHIV as compared to individuals without such illnesses (<u>19</u>, <u>33</u>, <u>34</u>, <u>36</u>, <u>40</u>). Opportunistic infections were substantially linked with undernutrition in PLHIV, with a combined odds ratio (OR) of 3.496 (95% CI 1.776, 5.217) when compared to those without these disorders. This finding shows that PLHIV with one or more OIs were around four times as likely to experience undernutrition than those without OIs.

Four studies found that WHO clinical stages II and above were related to undernutrition in PLHIV, relative to clinical stage I (<u>19</u>, <u>33</u>, <u>34</u>, <u>40</u>). The combined odds ratio (OR) for WHO clinical stages II and above in predicting undernutrition among PLHIV was 2.916 (95% confidence interval [CI] 1.088, 4.744) as compared to WHO clinical stage I. This finding suggests that PLHIV in Ethiopia with WHO clinical stage II or above were around three times more likely to be related to undernutrition than those at WHO clinical stage I.

In five of eleven studies, PLHIV with a CD_4_ level below 200 cells/mm³ were significantly connected to undernutrition compared to those with a CD_4_ count above 500 cells/mm³ (<u>19</u>, <u>22</u>, <u>33</u>, <u>36</u>, <u>39</u>). In five studies, the pooled odds ratio for undernutrition among PLHIV with a CD_4_ count less than 200 cells/mm³ was 3.099 (95% CI 1.418, 4.779), showing a higher risk than those with a CD_4_ count of 200 cells/mm³ and more (I² = 0.0%; p-value = 0.619). People living with HIV with a CD_4_ count <200 cells/mm³ were 3.099 times more likely to be undernourished than those with higher CD_4_ counts.

Food insecurity is another factor contributing to Ethiopia’s highest prevalence of undernutrition among PLHIV. In five searches, the pooled odds ratio for undernutrition among PLHIV with food insecurity was 3.625 (95% CI 1.460, 5.789), indicating a higher risk than their counterparts (I² = 0.0%; p-value = 0.983). People living with HIV who experienced food insecurity were 3.625 times more likely to be malnourished than those who were food secure ( <u>S5 Figure 1_4</u> and Table 6).

**Table 6:**
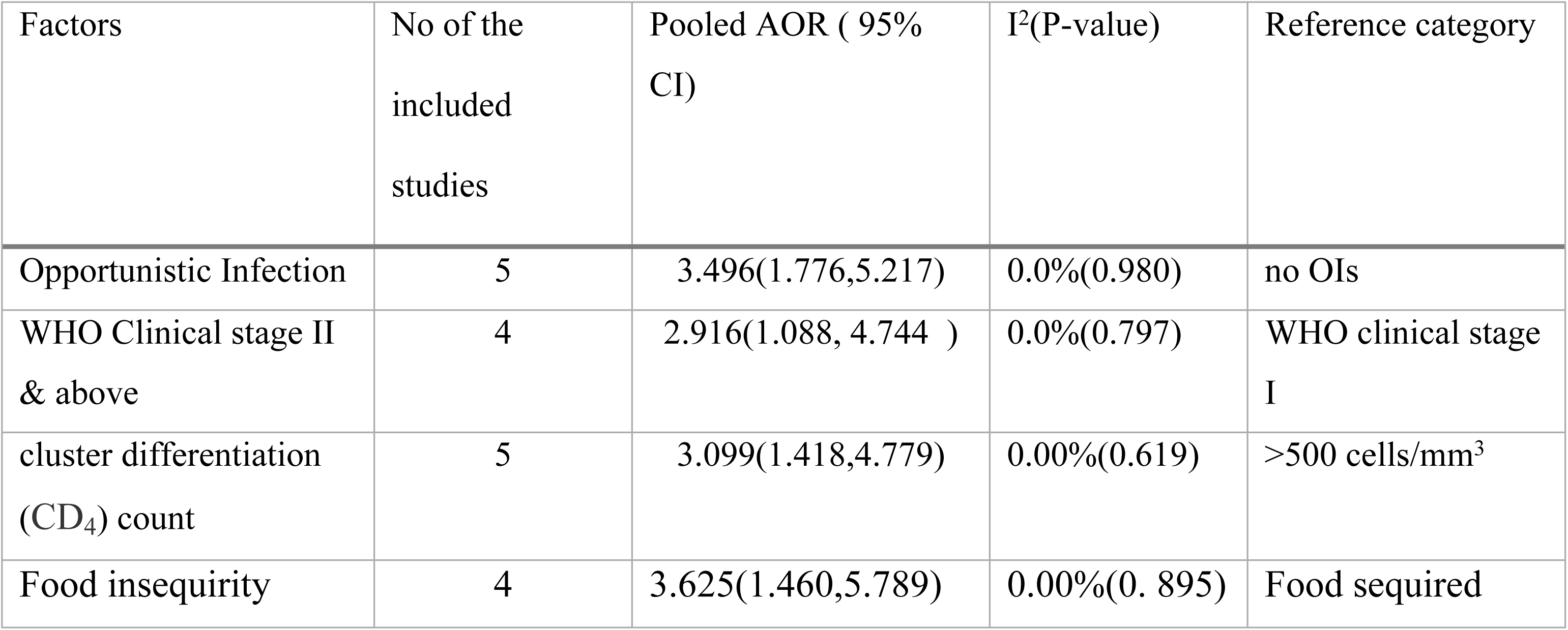
Summary of the factors associated with undernutrition among adults living with HIV in Ethiopia, 2023.

## 4. Discussion

The major goal of this study was to conduct an updated evidence synthesis of the prevalence and determining factors of undernutrition among PLHIV using a systematic review of discovering, selecting, and critically evaluating literature. As far as we know, this study is altering our knowledge based on earlier research findings. According to recent systematic reviews and meta-analyses conducted in Ethiopia, the prevalence of undernutrition is growing. While it is important to update knowledge and build upon previous research, it is possible that the prevalence of undernutrition is growing due to different methodologies or factors specific to the Ethiopian context. It is essential to consider these potential discrepancies before drawing definitive conclusions.

The study discovered that 26.70% of people with PLHIV in Ethiopia were malnourished, with a 95% confidence interval of 21.31% to 32.10%. Globally, undernutrition is linked to drought and climate change, both of which have a high positive correlation (<u>41</u>). The prevalence of undernutrition among PLHIV in the study was lower than in India, where it was 32.6% (<u>42</u>). This disparity could be explained by social and cultural differences. In comparison, the prevalence of undernutrition among PLHIV was greater than the 23.72% reported by the SSA (<u>43</u>). This disparity demonstrates that the highest prevalence of undernutrition among PLHIV is due to deep poverty, as Ethiopia is the lowest-income and most densely populated region in Africa and the world.

Furthermore, the findings of this study were consistent with a meta-analysis showing a 26% prevalence of undernutrition among PLHIV in Ethiopia (<u>24</u>). In addition, a systematic review and meta-analysis in the country found that the prevalence of undernutrition among PLHIV in Ethiopia has increased to 27.4%, which is greater than the current figure (<u>44</u>). Correlations do not imply causation, and comparing prevalence data may misrepresent Ethiopia’s condition. Future studies should look into the particular factors that contribute to undernutrition in PLHIV, and interventions should take into account the complex interaction of determinants in order to be effective and achieve desired health outcomes.

Subgroup analysis revealed significant variation in the prevalence of undernutrition among PLHIV across Ethiopia. It ranged from 16.0% to 42.90%, with the lowest prevalence reported in the SNNPR region and the highest in Tigray. Undernutrition prevalence varies between Ethiopian regions, which could be attributed to differences in socioeconomic level and food security (<u>45</u>, <u>46</u>). The SNNPR has faced substantial sociodemographic and economic challenges, resulting in acute food insecurity relative to other Ethiopian regions (<u>47</u>). This leads to an increased intake of carbohydrate-rich basic foods. As a result, malnutrition and other health conditions became more prevalent. Their society regards overweight people as a symbol of riches rather than a health risk. Thus, the low prevalence of undernutrition may be misconstrued.

In three SNNPR studies, the prevalence of undernutrition was 23.30% (95% CI: 14.70, 31.90), while in two Amhara studies, it was 28.32% (95% CI: 11.07, 45.57) (Fig. 3). One study indicated that the prevalence of undernutrition in the Harari regional state was 19.10% (95% CI: 15.66, 22.54), whereas in the Tigray regional state it was 42.90% (95% CI: 38.01, 47.79). It is important to consider the limitations of cross-sectional studies, as they may not capture the full picture of undernutrition prevalence over time or account for various factors that could influence the results. Additionally, regional differences in undernutrition prevalence could be influenced by a variety of factors such as access to healthcare, food security, and socio-economic status.

Our systematic review and meta-analysis revealed that opportunistic infections were associated with a higher frequency of undernutrition among adult PLHIV. Opportunistic infections were significantly associated with undernutrition in PLHIV, with a pooled odds ratio of 3.496 (95% CI 1.776, 5.217) compared to individuals without these illnesses (Table 6). At the same time, another study discovered that people with undernutrition are more likely to get OIs (<u>48</u>). A study conducted in Brazil and SSA found that OIs in adults with PLHIV increase the risk of undernutrition and decrease treatment outcomes due to reduced immunity (<u>49</u>, <u>50</u>). Adults with PLHIV who have previously been exposed to OIs in low- and middle-income countries have a higher risk of malnutrition (<u>51</u>). It is also the leading cause of death among HIV-positive adults (<u>52</u>). Thus, OIs and undernutrition in PLHIV on ART are strongly linked. While it is true that OIs can increase the risk of undernutrition in PLHIV, it is important to note that proper nutrition can also help improve immunity and decrease the likelihood of developing OIs. Additionally, advancements in antiretroviral therapy have significantly improved treatment outcomes for PLHIV, reducing the impact of OIs on overall health.

The advanced WHO clinical stage was identified as a determinant factor for the higher prevalence of undernutrition in Ethiopia among adults with PLHIV (Table 6). The combined odds ratio for WHO clinical stages II and above in predicting undernutrition among PLHIV was 2.916 (95% CI 1.088, 4.744) as compared to WHO clinical stage I. It agrees with the discoveries from the meta-analysis results in SSA and Ethiopia (<u>24</u>, <u>43</u>). Similarly, in Zimbabwe, undernutrition was 2.25 times more likely to be associated with an advanced WHO clinical stage of the disease (<u>16</u>). Thus, care for the undernourished adults in PLHIV cases in Ethiopia is high because undernutrition contributes to an increased risk of mortality and other adverse health outcomes (<u>50</u>). Advanced WHO clinical stages may contribute to undernutrition in Ethiopian PLHIV patients, but other factors and individual dietary requirements must also be considered.

Determinant factors of undernutrition in Ethiopia among adults with PLHIV were identified as having a low CD_4_ count (<200 cells/mm3). The pooled odds ratio for undernutrition among PLHIV with a CD_4_ count less than 200 cells/mm³ was 3.099 (95% CI 1.418, 4.779), showing a higher risk than those with a CD_4_count of 200 cells/mm³ and more (I² = 0.0%; p-value = 0.619). It matches the findings from the meta-analyses from 2006 to 2016 and the nationwide meta-analysis in Ethiopia (<u>24</u>). Similarly, a systematic review and meta-analysis (SRMA) conducted in SSA reported that a low CD_4_ count (<200 cells/mm3) was 1.94 times more associated with undernutrition among adults with PLHIV (<u>43</u>). The biological link between disease stages and undernutrition could be explained by the fact that as the disease progresses, the concurrence and recurrence of opportunistic infections worsen. This may be due to food insecurity, which could be a potential barrier to immune recovery as measured by CD_4_ counts among HIV-infected people (<u>53</u>). This may realize that undernutrition has been linked to HIV-related immune status, resulting in a low CD_4_count and possibly necessitating treatment at ART sites in Ethiopia due to exacerbated immune system decline.

Food instability is another factor contributing to Ethiopia’s high prevalence of undernutrition among people living with HIV. It was discovered that PLHIV experiencing food insecurity were 3.625 times more likely to be undernourished than those who were food secure, indicating a significantly greater risk (95% CI 1.460, 5.789) (Table 6). The relationship between undernutrition, access to food, and the health outcomes of HIV clients in Ethiopia is crucial. Implementing targeted interventions like food assistance programs and agricultural support can help alleviate undernutrition and improve overall health (<u>S5 Figue A-D</u>).

### 4.1. Implications of the findings

The current study varies from prior systematic reviews and meta-analyses in that it assessed the combined probability ratio of the factors influencing undernutrition among PLHIV in Ethiopia. It contains up-to-date findings and is more thorough because it was evaluated at the national level using substantial data. It could help government officials, policymakers, researchers, and other stakeholders assess the overall prevalence of undernutrition in Ethiopia’s various regions. Furthermore, allocate resources appropriately to address the deciding factors substantially associated with undernutrition in the study groups nationwide. The study focuses on undernutrition among PLHIV in Ethiopia, but it suggests a comprehensive approach that considers social, economic, and environmental aspects in order to conduct effective interventions.

## 5. Conclusion

According to the evaluated publications, undernutrition among individuals with HIV is a notable public health concern in Ethiopia. Exposure to opportunistic infections, an advanced disease stage (II-V), and a CD_4_ count of less than 200 cells/mm3 were found as major risk factors for undernutrition among people with PLHIV in Ethiopia. Adults with HIV who are experiencing these diseases require additional attention because they have been identified as major contributors to Ethiopia’s growing prevalence of undernutrition. Health is threatened not only by disease and infections, but also by underlying causes such as poverty and food insecurity, which determine the frequency of health issues and the interplay between them. Further research could look into how opportunistic infections, advanced WHO clinical stages, and a CD_4_ count below 200 cells/mm³ impact the frequency of undernutrition among HIV-positive individuals.

## Data Availability

All data are within the manuscript and its supporting information files.

## Ethical approval and consent for participation

It is not applicable.

## Consent for publication

It is not applicable.

## Availability of data and materials

All required data used for the analysis will be available from the corresponding author based on requests from reviewers.

## Complict of interest

All authors declered that they had no competing interests.

## Authors contributions

Amare Admasu (A.A) has drafted the work, designed the work, screened it, reviewed it, performed quality checks, performed result analysis, interpreted the data, substantively revised it, and approved the submitted version.

Takeke Tadese (TT), Amene Abebe (AA), and Eskinder Wolka (EW): integrity of any part of the work, screening, reviewing, quality checks, result analysis, investigation, resolution, and the resolution documented in the SRMA, and have approved the submitted version. All authors read and approved the final manuscript.

## Acknowledgment

First and foremost, I would like to thank Wolaita Sodo University for giving me this chance to follow this study. Then my thanks go to the assistance received and to those to whom he or she remains thankful for special aid or support.

## Authors information

1. Amare Admasu (Candidate for a PhD in public health, MSC in human nutrition, clinician at Wolaita Sodo University comprehensive specialized hospital, Woliata, Ethiopia)
2. Professor Takele Tadesse (Ph.D., Prof. Epidemiology and Biostatistics, lecture in Wolaita Sodo University College of Health Science and Medicine, Wolaita, Ethiopia)
3. Amene Abebe (Ph.D., Associate Professor of Public Health, lecture in Wolaita Sodo University College of Health Science and Medicine, Wolaita, Ethiopia)
4. Eskindir Wolka (Ph.D., Assistant Professor of Public Health, lecture in Wolaita Sodo University College of Health Science and Medicine, Wolaita, Ethiopia)

## Abbreviations/Acronyms

PLHIV: People Living with HIV
SRMA: systematic review and meta-analysis
SSA: Sub-Saharan-Africa
WLH: women living with HIV

---

**A: Figure l:**
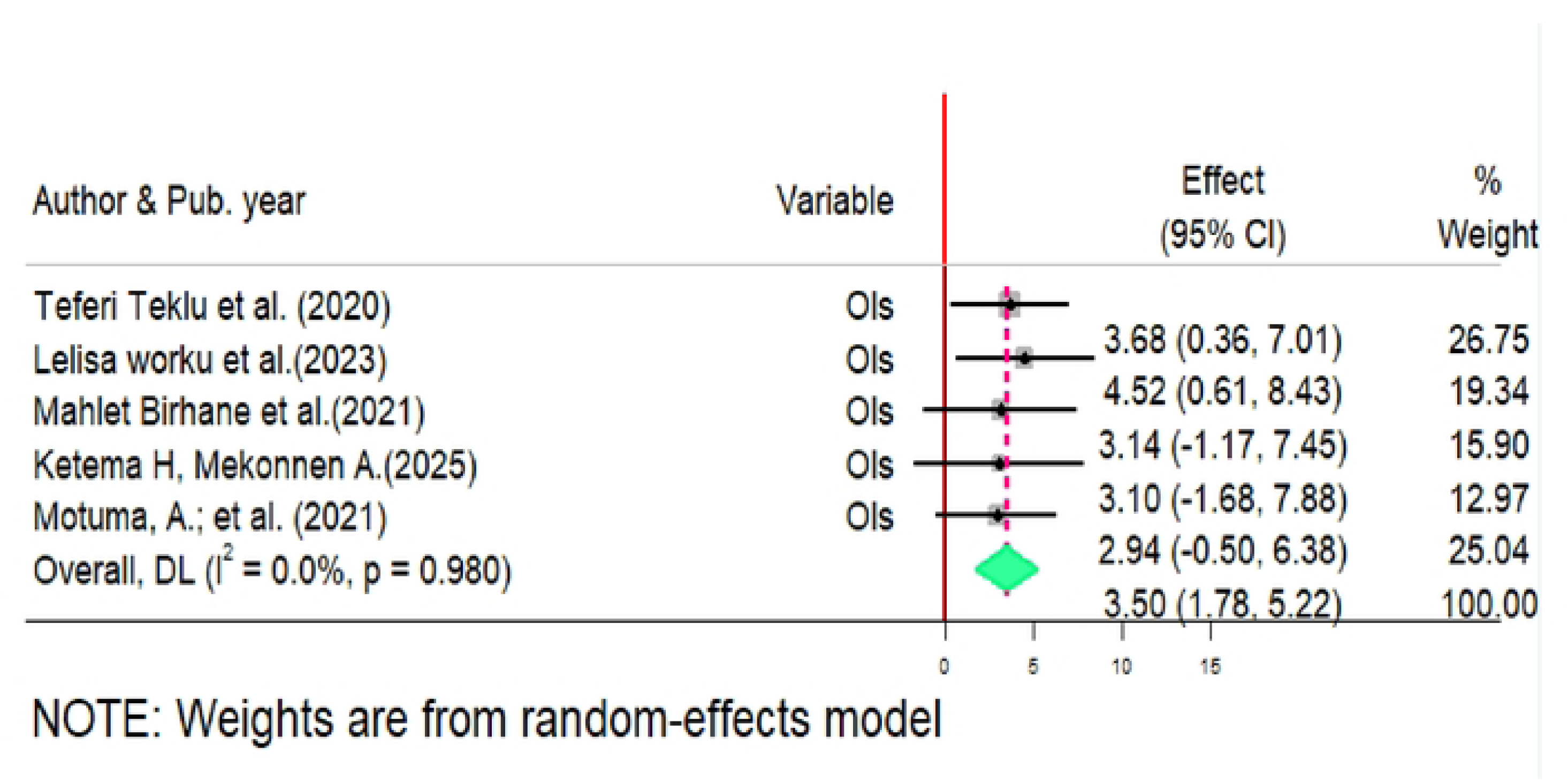
Forest plot of opportunistic infections (OIs) as determinant factors among PLHIV in Ethiopia from 2020 to 2025.

**B: Figure 2:**
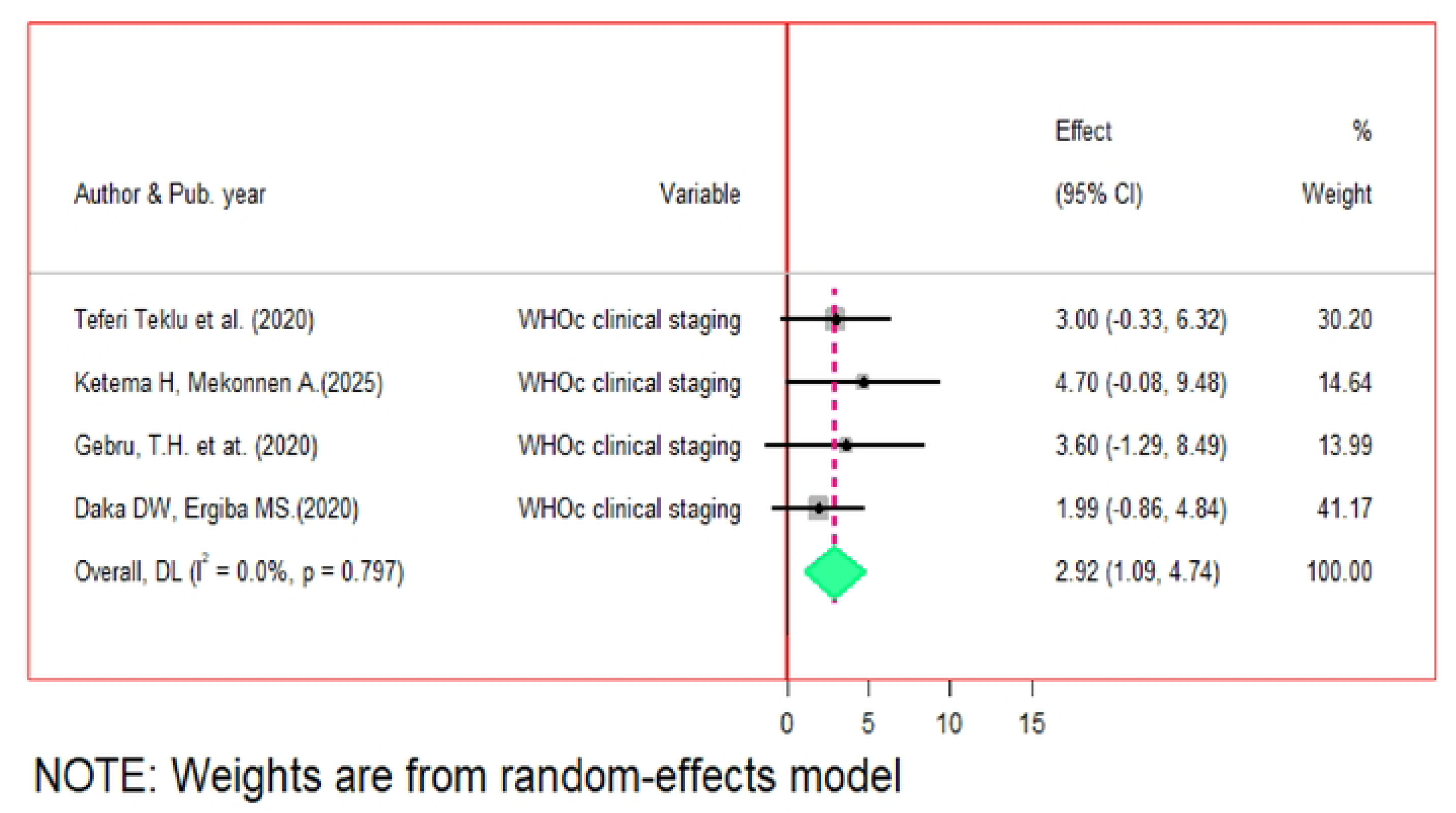
Forest plot of advanced WHO clinical stage as a determinant factors among PLRIV in Ethiopia from 2020 to 2025.

**C: Figure 3:**
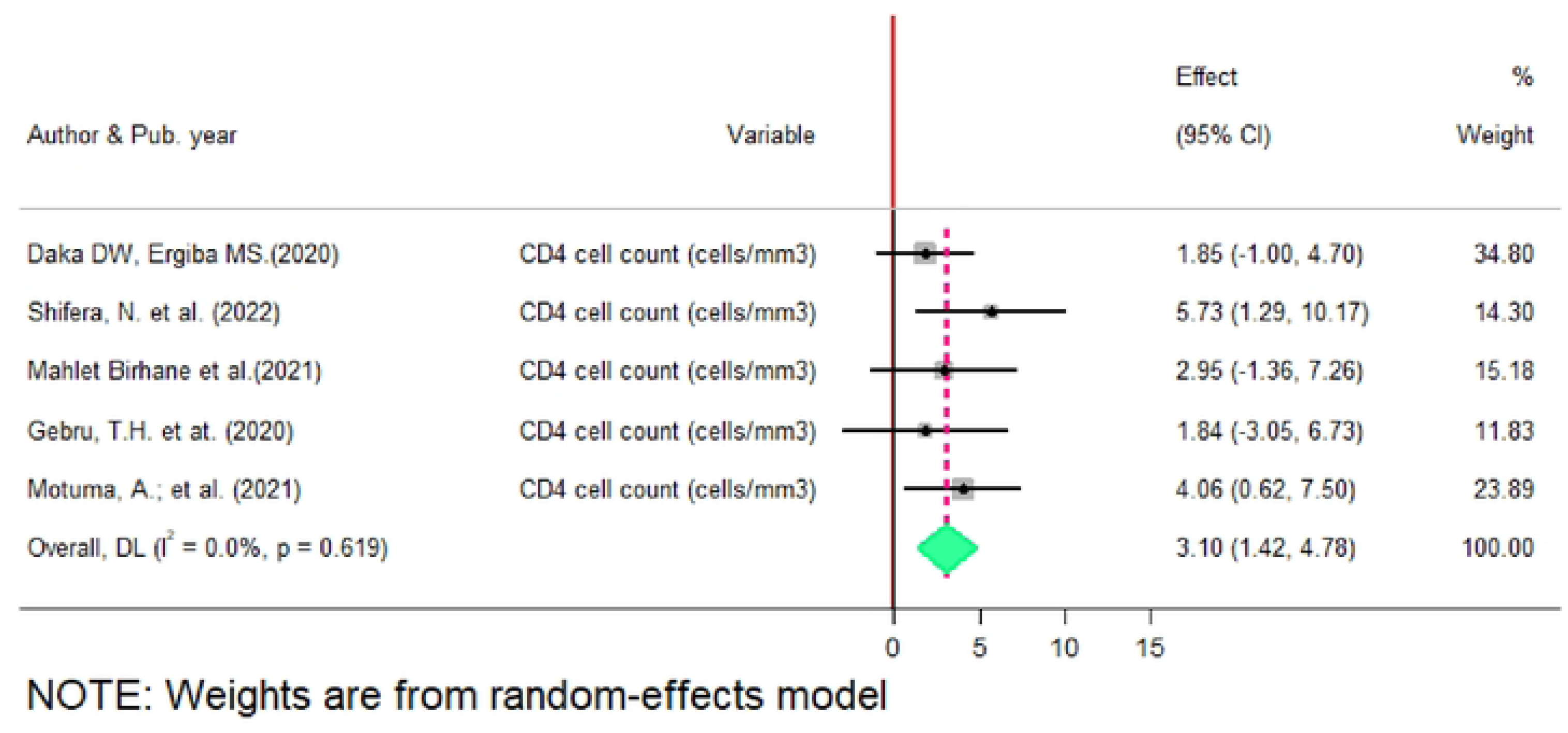
Forest plot of CD_4_ count as determinant factors among PLHIV in Ethiopia from 2020 to 2025.

**D: Figure 4:**
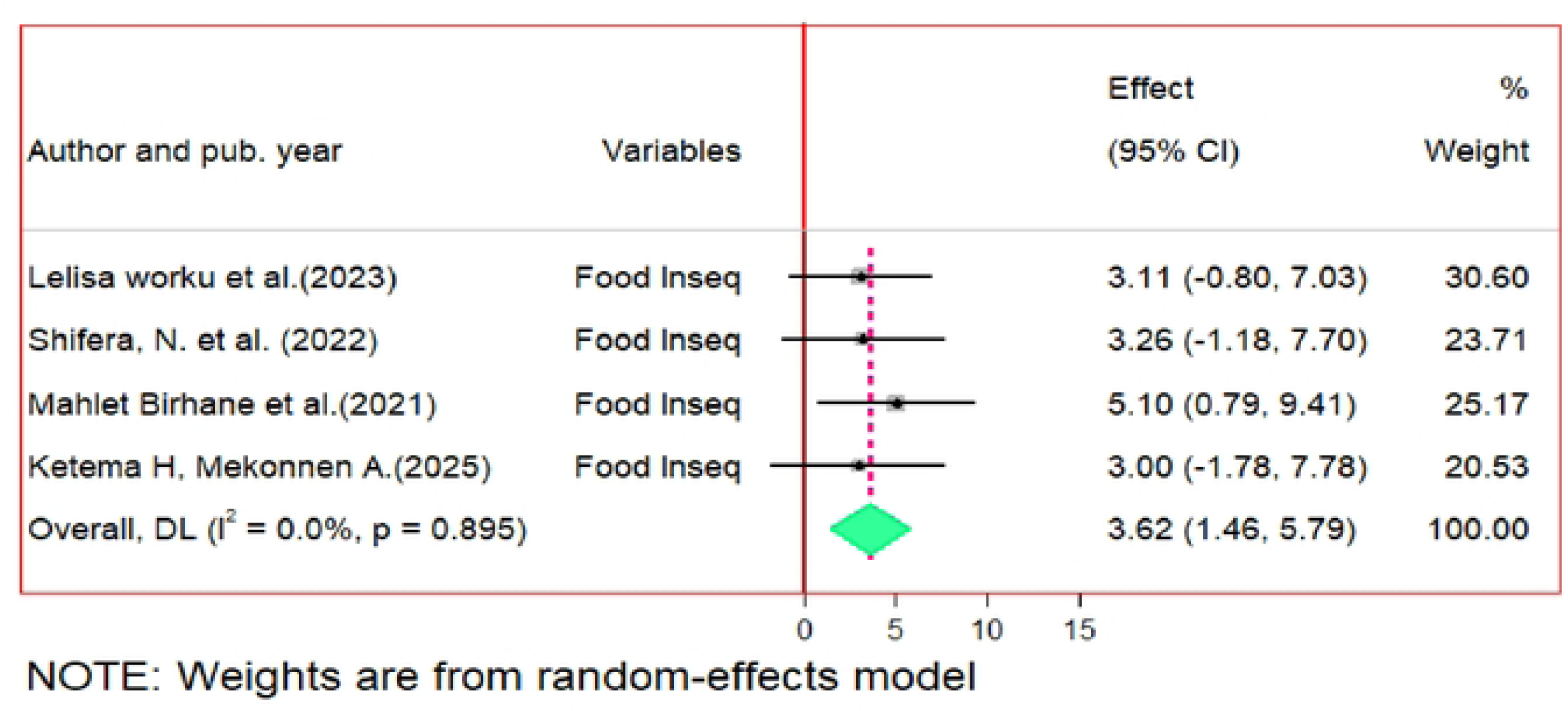
Forest plot of food insecurity as a determinant factors among PLRIV in Ethiopia from 2020 to 2025.

## References

1. FAO. Minimum Dietary Diversity for Women- A Guide to Measurement. 10.4060/cb3434en. Food and Agriculture Organization of the United Nations Rome: 2021. p. 82.

2. Government of Ethiopia, Federal Ministry of health. National Guideline for the Management of Acute Malnutrition in Ethiopia. In: FMoH, editor. Addis Ababa, Ethiopia. 2019.

3. Hinke Kruizenga PhD RD, Sandra Beijer PhD RD, Getty Huisman-de Waal PhD RN, Cora Jonkers-Schuitema Bsc RD, Mariël Klos Bsc RN, Wineke Remijnse-Meester Bsc RD, et al. Guideline on malnutrition. Recognising, diagnosing and treating malnutrition in adults Dutch Malnutrition Steering Group. 2017. 2017.

4. World Health Organization. Antiretroviral therapy for HIV infection in adults and adolescents: recommendations for a public health approach-2010 revision: https://iris.who.int/bitstream/handle/10665/44379/9789241599764_eng.pdf?sequence=12010.

5. IPC Global Partners. Integrated Food Security Phase Classification Technical Manual Version 3.1. Evidence and Standards for Better Food Security and Nutrition Decisions. https://www.ipcinfo.org/fileadmin/user_upload/ipcinfo/manual/IPC_Technical_Manual_3_Final.pdf. Rome2021.

6. Definitions, Classification, and Epidemiology of Obesity. 2023 May 4. In: Feingold KR, Anawalt B, Blackman MR, Boyce A, Chrousos G, Corpas E, de Herder WW, Dhatariya K, Dungan K, Hofland J, Kalra S, Kaltsas G, Kapoor N, Koch C, Kopp P, Korbonits M, Kovacs CS, Kuohung W, Laferrère B, Levy M, McGee EA, McLachlan R, New M, Purnell J, Sahay R, Shah AS, Singer F, Sperling MA, Stratakis CA, Trence DL, Wilson DP, editors. Endotext [Internet]. South Dartmouth (MA): MDText.com, Inc.; 2000–. PMID: 25905390. [Internet]. 2023. Available from: http://creativecommons.org/licenses/by-nc-nd/2.0/.

7. UNAIDS. FACT SHEET 2023: Global HIV statistics.https://www.unaids.org/sites/default/files/media_asset/UNAIDS_FactSheet_en.pdf. 2023.

8. Jennifer Peregoy. HIV/AIDS, UNDER-NUTRITION AND FOOD INSECURITY. https://www.worldhunger.org/hivaids-under-nutrition-and-food-insecurity-2/. 2017.

9. World Health Organization. The state of food security and nutrition in the world 2019: safeguarding against economic slowdowns and downturns: Food & Agriculture Organization: https://www.wfp.org/publications/2019-state-food-security-and-nutrition-world-sofi-safeguarding-against-economic. 2019.

10. Organization WH. Global Health Observatory (GHO) data. Geneva: WHO; 2020 [cited 12 May 2020]. https://www.who.int/data/gho/data/themes/hiv-aids.

11. Folasire O, Folasire A, Sanusi R. Measures of nutritional status and quality of life in adult people living with HIV/AIDS at a tertiary hospital in Nigeria. Food and Nutrition Sciences. 2015;6(04):412.

12. Kabalimu TK, Sungwa E, Lwabukuna WC. Malnutrition and associated factors among adults starting on antiretroviral therapy at PASADA Hospital in Temeke District, Tanzania. Tanzania Journal of Health Research. 2018;20(2).

13. Benzekri NA, Sambou J, Diaw B, Sall EHI, Sall F, Niang A, et al. High prevalence of severe food insecurity and malnutrition among HIV-infected adults in Senegal, West Africa. PloS one. 2015;10(11):e0141819.

14. Nanewortor BM, Saah FI, Appiah PK, Amu H, Kissah-Korsah K. Nutritional status and associated factors among people living with HIV/AIDS in Ghana: cross-sectional study of highly active antiretroviral therapy clients. BMC nutrition. 2021;7(1):1–8.

15. Odwee A, Kasozi KI, Acup CA, Kyamanywa P, Ssebuufu R, Obura R, et al. Malnutrition amongst HIV adult patients in selected hospitals of Bushenyi district in southwestern Uganda. African health sciences. 2020;20(1):122–31.

16. Takarinda KC, Mutasa-Apollo T, Madzima B, Nkomo B, Chigumira A, Banda M, et al. Malnutrition status and associated factors among HIV-positive patients enrolled in ART clinics in Zimbabwe. BMC Nutrition. 2017;3:1–11.

17. Daniel M, Mazengia F, Birhanu D. Nutritional status and associated factors among adult HIV/AIDS clients in Felege Hiwot Referral Hospital, Bahir Dar, Ethiopia. Science Journal of Public Health. 2013;1(1):24–31.

18. Alebel A, Demant D, Petrucka P, Sibbritt D. Effects of undernutrition on opportunistic infections among adults living with HIV on ART in Northwest Ethiopia: Using inverse-probability weighting. Plos one. 2022;17(3):e0264843.

19. Daka DW, Ergiba MS. Prevalence of malnutrition and associated factors among adult patients on antiretroviral therapy follow-up care in Jimma Medical Center, Southwest Ethiopia. PLoS One. 2020;15(3):e0229883.

20. Dedha M, Damena M, Egata G, Negesa L. Undernutrition and associated factors among adults human immunodeficiency virus positive on antiretroviral therapy in hospitals, East Hararge Zone, Oromia, Ethiopia: a cross-sectional study. International journal of health sciences. 2017;11(5):35.

21. Nigusso FT, Mavhandu-Mudzusi AH. High magnitude of food insecurity and malnutrition among people living with HIV/AIDS in Ethiopia: A call for integration of food and nutrition security with HIV treatment and care Programme. Nutrition and health. 2021;27(2):141–50.

22. Birhane M, Loha E, Alemayehu FR. Nutritional status and associated factors among adult HIV/AIDS patients receiving ART in Dilla University referral hospital, Dilla, Southern Ethiopia. J Med Physiol Biophys. 2021;70:8–15.

23. Asnakew M. Malnutrition and associated factors among adult individuals receiving highly active antiretroviral therapy in health facilities of Hosanna Town, Southern Ethiopia. Open Access Library Journal. 2015;2(01):1.

24. Alebel A, Kibret GD, Petrucka P, Tesema C, Moges NA, Wagnew F, et al. Undernutrition among Ethiopian adults living with HIV: a meta-analysis. BMC nutrition. 2020;6(1):1–10.

25. Page MJ, McKenzie JE, Bossuyt PM, Boutron I, Hoffmann TC, Mulrow CD, et al. The PRISMA 2020 statement: an updated guideline for reporting systematic reviews. International journal of surgery. 2021;88:105906.

26. Grainge M, editor Excluding small studies from a systematic review or meta-analysis. CSG Annual Meeting; 2015.

27. Peterson J, Welch V, Losos M, Tugwell P. The Newcastle-Ottawa scale (NOS) for assessing the quality of nonrandomised studies in meta-analyses. Ottawa: Ottawa Hospital Research Institute. 2011;2(1):1–12.

28. Organization WH. Nutrition Landscape Information System (NLIS) country profile indicators: interpretation guide. 2019.

29. Peters MD, Godfrey CM, McInerney P, Soares CB, Khalil H, Parker D. The Joanna Briggs Institute reviewers’ manual 2015: methodology for JBI scoping reviews. 2015.

30. Higgins JP, Thompson SG, Deeks JJ, Altman DG. Measuring inconsistency in meta-analyses. bmj. 2003;327(7414):557–60.

31. Rücker G, Schwarzer G, Carpenter JR, Schumacher M. Undue reliance on I 2 in assessing heterogeneity may mislead. BMC medical research methodology. 2008;8:1–9.

32. Fantahun Guteta, Gemechu Kejela. Under Nutrition and Associated Factors among Adult HIV/AIDS Patients Enrolled on ART at Public Health Facilities of Western Ethiopia. . Virology & Mycology. 2021;20(5):1–6.

33. Gebru TH, Mekonen HH, Kiros KG. Undernutrition and associated factors among adult HIV/AIDS patients receiving antiretroviral therapy in eastern zone of Tigray, Northern Ethiopia: a cross-sectional study. Archives of public health = Archives belges de sante publique. 2020;78:100.

34. Ketema H, Mekonnen A. Nutritional Status and associated factors among adults living with HIV/AIDS in Yekatit 12 Hospital, Addis Ababa, Ethiopia: A facility-based cross-sectional study. available at: https://mjh.sphmmc.edu.et/volume_IV_issue_I/MJH-2024-0024.published.pdf. 4. 2024;1:2790–1378.

35. Lelisa worku, Dube Jara, Tola Abera, Eyerusalem Degife, Gadise Regassa, Sisay Hailu. Undernutrition and Associated Factors among HIV-Infected Adults Receiving ART at Health Centers, Lega Tafo and Surrounding, Ethiopia, 2021. International Journal of Clinical and Medical Education Research. 2023;2(4):89–96.

36. Motuma A, Abdeta T. Undernutrition and Associated Factors Among Seropositive Adults in ART Clinic Treatment Centre, Hiwot Fana Specialized University Hospital, Eastern Ethiopia. Pathology and Laboratory Medicine. 2021;5(1):10.

37. Regassa TM, Gudeta TA. Levels of undernutrition and associated factors among adults receiving highly active anti-retroviral therapy in health institutions in Bench Maji Zone, Southwest Ethiopia in 2018. Frontiers in nutrition. 2022;9:814494.

38. Sahile AT, Ayehu SM, Fanta SF. Underweight and Its Predictors Among Patients on Anti Retroviral Therapy at Selected Health Facilities of Addis Ababa, Ethiopia, 2020. HIV/AIDS-Research and Palliative Care. 2021:99–106.

39. Shifera N, Yosef T, Matiyas R, Kassie A, Assefa A, Molla A. Undernutrition and Associated Risk Factors among Adult HIV/AIDS Patients Attending Antiretroviral Therapy at Public Hospitals of Bench Sheko Zone, Southwest Ethiopia. Journal of the International Association of Providers of AIDS Care. 2022;21:23259582221079154.

40. Teklu T, Chauhan NM, Lemessa F, Teshome G. Assessment of Prevalence of Malnutrition and Its Associated Factors among AIDS Patients from Asella, Oromia, Ethiopia. Biomed Res Int. 2020;2020:7360190.

41. Lieber M, Chin-Hong P, Kelly K, Dandu M, Weiser S. A systematic review and meta-analysis assessing the impact of droughts, flooding, and climate variability on malnutrition. Global Public Health. 2020;17:1–15.

42. Parida J, Bagepally BS, Patra PK, Pati S, Kaur H, Acharya SK. Prevalence and associated factors of undernutrition among adolescents in India: a systematic review. BMC public health. 2025;25(1):819.

43. Seid A, Seid O, Workineh Y, Dessie G, Bitew ZW. Prevalence of undernutrition and associated factors among adults taking antiretroviral therapy in sub-Saharan Africa: A systematic review and meta-analysis. : e0283502. 10.1371/journal.pone.0283502. PLoS ONE. 2023;18(3).

44. Mengie T, Dejen D, Muche T, Getacher L, Kindie B, Tamiru C. Under Nutrition and Its Determinants Among Adults Receiving Antiretroviral Therapy in Ethiopia: A Systematic Review and Meta-analysis. Int J Homeopathy Nat Med. 2021;7(1):1–6.

45. Boneya DJ, Ahmed AA, Yalew AW. The effect of gender on food insecurity among HIV-infected people receiving anti-retroviral therapy: A systematic review and meta-analysis. PLoS One. 2019;14(1):e0209903.

46. Demisse A, Demena M, Ayele BH, Mengistu A. Food insecurity and associated factors among adult HIV patients on anti-retroviral therapy in Dessie referral hospital, South Wollo Zone, North central Ethiopia. PLOS Global Public Health. 2022;2(9):e0000445.

47. Ayana Anteneh Astatike, Ermias Ganamo Gazuma, Tatasha Takaye Kasito. Rural poverty and its determinants in wolaita zone, southern Ethiopia. International Journal of Development Research. 2019;09(05):27972–7.

48. Alebel A, Demant D, Petrucka P, Sibbritt D. Effects of undernutrition on mortality and morbidity among adults living with HIV in sub-Saharan Africa: a systematic review and meta-analysis. BMC infectious diseases. 2021;21(1):1–20.

49. Bassichetto KC, Bergamaschi DP, Garcia VRS, Veras MAdSM. Factors associated with undernourishment among people 20 years old or over with HIV/AIDS, attending public health services in the São Paulo municipality, Brazil. Cadernos de Saúde Pública. 2014;30:2578–86.

50. Fuseini H, Gyan BA, Kyei GB, Heimburger DC, Koethe JR. Undernutrition and HIV infection in sub-Saharan Africa: health outcomes and therapeutic interventions. Current HIV/AIDS Reports. 2021;18:87–97.

51. Gebrie M, Perry L, Xu X, Kassa A, Cruickshank M. Nutritional status and its determinants among adolescents with HIV on anti-retroviral treatment in low-and middle-income countries: a systematic review and meta-analysis. BMC nutrition. 2023;9(1):60.

52. Misgina KH, Weldu MG, Gebremariam TH, Weledehaweria NB, Alema HB, Gebregiorgis YS, et al. Predictors of mortality among adult people living with HIV/AIDS on antiretroviral therapy at Suhul Hospital, Tigrai, Northern Ethiopia: a retrospective follow-up study. Journal of Health, Population and Nutrition. 2019;38:1–10.

53. Aibibula W, Cox J, Hamelin A-M, Mamiya H, Klein MB, Brassard P. Food insecurity and low CD4 count among HIV-infected people: a systematic review and meta-analysis. AIDS care. 2016;28(12):1577–85.

